# Molecular surveillance to monitor the prevalence of tetracycline resistance in *Neisseria gonorrhoeae*

**DOI:** 10.1101/2024.05.07.24306823

**Authors:** Kirstin I. Oliveira Roster, Rachel Mittelstaedt, Jordan Reyes, Aishani V. Aatresh, Yonatan H. Grad

## Abstract

Doxycycline post-exposure prophylaxis (Doxy-PEP) reduces bacterial sexually transmitted infections (STIs) but may select for tetracycline resistance in Neisseria gonorrhoeae and co-resistance to other antibiotics, including ceftriaxone.. The implementation of doxy-PEP should be accompanied by monitoring doxycycline resistance, but the optimal strategy to detect changes in the prevalence of resistance has not been established. We used a deterministic compartmental model of gonorrhea transmission to evaluate the performance of two strategies in providing early warning signals for rising resistance: (1) phenotypic testing of cultured isolates and (2) PCR for tetM in remnants from positive Nucleic Acid Amplification Tests (NAATs) used for gonorrhea diagnosis. For each strategy, we calculated the resistance proportion with 90% simulation intervals as well as the time under each sampling strategy to achieve 95% confidence that the resistance proportion exceeded a resistance threshold ranging from 11-30%. Given the substantially larger available sample size, PCR for tetM in remnant NAATs detected increased high-level tetracycline resistance with high confidence faster than phenotypic testing of cultured specimens. Our results suggest that population surveillance using molecular testing for tetM can complement culture-based surveillance of tetracycline resistance in N. gonorrhoeae and inform policy considerations for doxy-PEP.

---

Doxycycline post-exposure prophylaxis (doxy-PEP) reduces bacterial sexually transmitted infections (STIs) (1). However, doxy-PEP may select for tetracycline resistance in *Neisseria gonorrhoeae* and co-resistance to other antibiotics, including ceftriaxone (2,3). The implementation of doxy-PEP should be accompanied by monitoring doxycycline resistance, but the optimal strategy to detect changes in the prevalence of resistance has not been established.

We used a deterministic compartmental model of gonorrhea transmission (4) to evaluate the performance of two strategies in providing early warning signals for rising resistance. First, we considered culture-based phenotypic testing for tetracycline resistance, with sampling based on the CDC’s Gonococcal Isolate Surveillance Project (GISP) and Enhanced GISP (eGISP) (5). Second, we considered the use of a molecular assay that tests for the presence of *tetM* in remnants from positive Nucleic Acid Amplification Tests (NAATs), building on the expanding repertoire of molecular resistance tests (5–7). While the tetracycline MIC that confers doxy-PEP resistance is as yet unclear, minocycline pre-exposure prophylaxis for gonococcal urethritis failed with tetracycline MIC > 2μg/mL (8); as such, it is reasonable to expect that high-level tetracycline resistance (MIC > 8 μg/mL), conferred by *tetM* (2), will be selected by doxy-PEP.

We simulated transmission of tetracycline-resistant, ceftriaxone-resistant, dual resistant, and fully susceptible gonorrhea strains. Each month, we sampled 25 cultured specimens from all symptomatic urethral cases presenting to care and 20% of positive NAATs, regardless of symptom status. Sampling was simulated as a binomial process with 1000 iterations. In sensitivity analyses, we considered sampling intensities of 5-80 monthly cultured specimens and 5-80% of positive NAATs. For each surveillance strategy, we calculated the mean high-level tetracycline resistance proportion and 90% simulation intervals and compared them to the true resistance proportion. The primary study outcome was the time under each sampling strategy to achieve 95% confidence that the resistance proportion exceeded a resistance threshold ranging from 11-30% (**Supplemental Methods, Figure S1, Table S1**).

While both molecular and culture-based surveillance produced similar estimates of the high-level tetracycline resistance proportion, the two strategies differed considerably in the variation of their estimates (**Figure 1a**). Because of the smaller sample size, culture-based surveillance yielded wide confidence intervals around the estimated resistance proportion, thus prolonging the time required to confidently detect rising resistance. In the simulations, after 6 months of doxy-PEP use in 30% of the population, 19.3% of monthly infections were caused by strains with high-level tetracycline resistance. Molecular surveillance estimated the resistance proportion at 19.1% (17.1-21.3%, 90% simulation interval), while phenotypic surveillance estimated a resistance proportion of 18.8% with substantial uncertainty (8.0-32.0%, 90% simulation interval).

**Figure 1.**
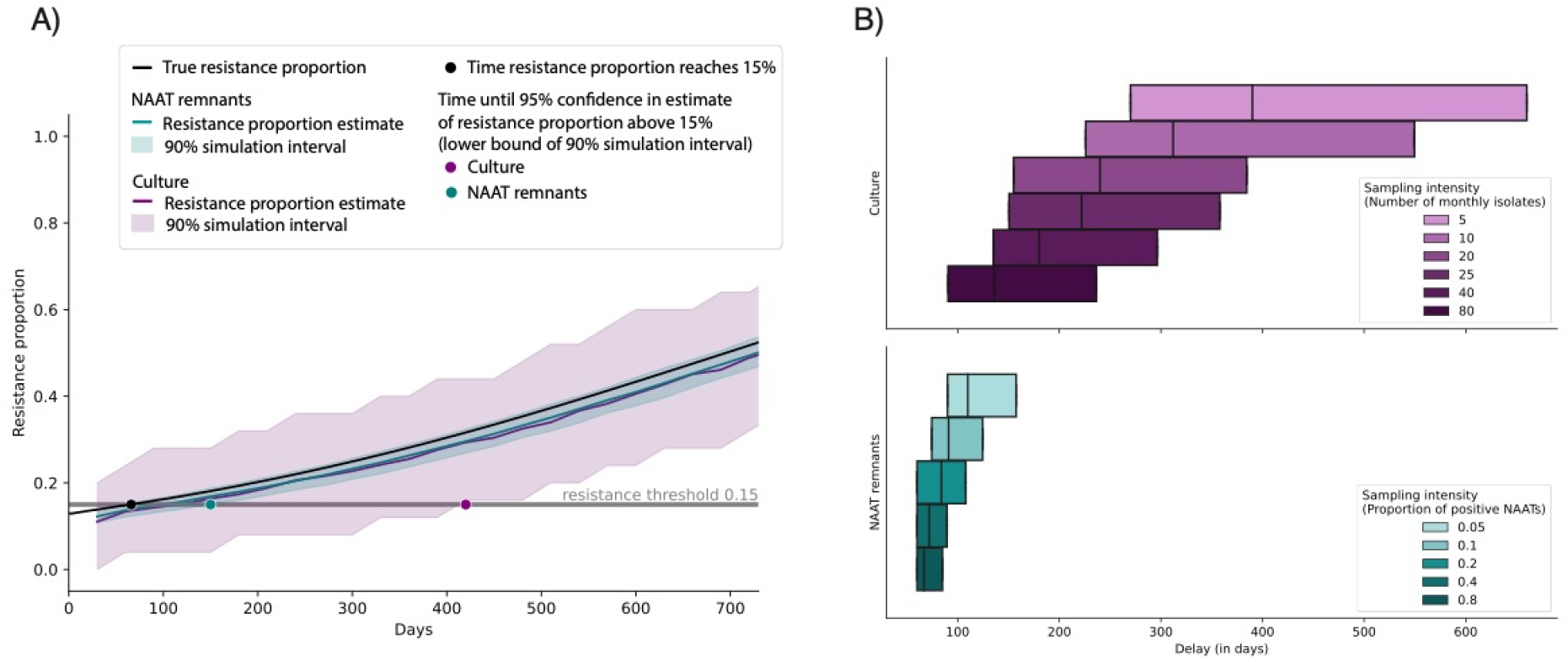
Estimates of tetracycline resistance from culture-based and molecular surveillance. **A**. Resistance proportion and 90% simulation intervals over time, estimated from 25 monthly cultured samples (purple) and 20% of positive NAATs (teal) relative to the true resistance proportion (black) under a doxy-PEP uptake rate of 30%. The black dot indicates the time the resistance proportion reaches a threshold of 15%. Colored dots indicate the time point at which each sampling method estimates a resistance proportion above the 15% threshold in 95% of simulations. **B**. Distributions (25^th^ percentile, median, and 75^th^ percentile) of the time delay until attaining 95% confidence in crossing a resistance threshold relative to the true time it takes for the resistance to reach the threshold, calculated over resistance thresholds ranging from 11-30% and doxy-PEP uptake levels from 10-90%. Sampling intensities range from 5-80 monthly cultured specimens (shades of purple) and 5-80% of positive NAATs (shades of teal).

We examined the time it took each strategy to accurately identify that resistance had crossed a given threshold in at least 95% of simulations. Under a scenario of 30% doxy-PEP uptake and 10.4% initial high-level tetracycline resistance (9), it took 66 days until 15% of infections were caused by resistant strains (**Figure 1a**). Molecular surveillance for *tetM* estimated resistance above the 15% threshold in 95% of simulations by 5 months (150 days), trailing the true resistance proportion by 84 days. With culture-based surveillance, 95% confidence was achieved after 14 months (420 days), lagging behind true resistance by 354 days.

For both strategies, increasing sampling improved the timeliness of confident early warning signals (**Figure 1b**). Scaling up phenotypic surveillance from 25 to 80 monthly cultured samples (from an average 0.66% to 2.1% of observed cases), reduced the median delay by 86 days (39%). Raising the proportion of sampled NAATs from 20 to 40% (from an average 747 to 1508 monthly samples), reduced the median delay by 12 days (14%). Results were consistent across doxy-PEP uptake scenarios (**Figure S2**).

Given the substantially larger sample size, molecular surveillance detected a rise in resistance earlier and with greater confidence compared to culture-based surveillance alone. Narrower confidence intervals meant that a reported increase in resistance was more likely to reflect a true rise in resistance and would thus enable a faster public health response. Our results suggest that molecular testing for *tetM* can complement culture-based surveillance of tetracycline resistance in *N. gonorrhoeae* and inform policy considerations for doxy-PEP.

## Data Availability

All code and data are available at: https://github.com/gradlab/doxy-PEP_surveillance

## Acknowledgments

The authors thank Samantha Palace for helpful comments on the manuscript.

The project described was supported by Grant Number T32 AI007433 from the National Institute of Allergy and Infectious Diseases. Its contents are solely the responsibility of the authors and do not necessarily represent the official views of the NIH.

This project has been funded (in part) by contract 200-2016-91779 with the Centers for Disease Control and Prevention. Disclaimer: The findings, conclusions, and views expressed are those of the author(s) and do not necessarily represent the official position of the Centers for Disease Control and Prevention (CDC).

