## Supplement for "Molecular surveillance to monitor the prevalence of tetracycline resistance in *Neisseria gonorrhoeae*"

### Supplementary Methods

In the United States, antibiotic susceptibility in *Neisseria gonorrhoeae* in the United States is monitored through the CDC's Gonococcal Isolate Surveillance Project (GISP), which performs phenotypic resistance testing for the first 25 symptomatic urethral infections diagnosed at sentinel sites across the country. Enhanced GISP (eGISP) tests up to 50 additional cultured specimens from extra-genital infections in men and from both genital and extra-genital infections in women at participating sentinel sites. For each of these infections, eGISP also collects remnants of nucleic acid amplification tests (NAATs), widely used for gonorrhea diagnosis, and conducts molecular testing for known resistance markers to four antibiotics, not yet including tetracycline (1).

Remnant NAAT surveillance could be expanded to test for the plasmid-encoded *tetM* gene, which confers high-level tetracycline resistance (with a minimum inhibitory concentration, MIC, above 8$\mu$g/mL) and is conservatively assumed to mediate doxy-PEP resistance. Molecular testing of NAAT remnants could complement culture-based approaches to scale up and broaden surveillance of *tetM*-mediated tetracycline resistance (2).

Using a deterministic compartmental model of gonorrhea (3), we simulated the transmission of tetracycline-resistant, ceftriaxone-resistant, dual resistant, and fully susceptible gonorrhea strains in a population of 1 million men who have sex with men (MSM), stratified by partner change rates into high, medium, and low risk groups. The initial prevalence of high-level tetracycline was 10.4%, consistent with reported levels in the US (4). The model considered doxy-PEP uptake rates of 10-90% and accounted for treatment with ceftriaxone in response to symptomatic care seeking and asymptomatic screening. In this model, infections initially dropped after rollout of doxy-PEP, followed by a rise in incidence due to selection for tetracycline-resistant strains. Higher doxy-PEP uptake led to not only a steeper initial decline in cases but also faster spread of resistance and therefore a faster uptick in cases after the initial dip (3). Model structure, equations, and parameters are listed below (**Figure S1, Table S1**). Further details are provided in Reichert and Grad (2023) (3).

We expanded this model to compare the ability to detect increases in tetracycline resistance in two approaches: (1) phenotypic testing of cultured isolates and (2) PCR for *tetM* in remnants from NAATs.

For phenotypic surveillance, we sampled 25 monthly cultured specimens from all symptomatic urethral cases presenting for care, consistent with the levels monitored as part of GISP. For molecular surveillance, we sampled 20% of positive NAATs, regardless of symptom status. In sensitivity analyses, we considered sampling intensities of 5-80 monthly cultured specimens and 5-80% of positive NAATs. Sampling was simulated as a binomial process with 1000 iterations.

For each simulation, we calculated the proportion of samples that were tetracycline resistant at each sampling time point. Cultured specimens were tested monthly. NAATs were sampled daily and pooled to generate monthly resistance proportion estimates.

We then calculated the mean tetracycline resistance proportion and 90% simulation intervals across all simulations. We compared these estimates with the true resistance proportion of all infections in the population. The primary study outcome was the time it would take under each sampling strategy to be 95% confident that the resistance proportion exceeded a resistance threshold ranging from 11-30%.

All code is available at https://github.com/gradlab/doxy-PEP_surveillance.

### Limitations

The model in this study accounted for the key differences between molecular and culture-based surveillance but took a simplified perspective of surveillance overall. For example, we assumed population-wide random sampling for both strategies. Non-random sampling - for example, due to geographic and socioeconomic differences in care seeking behavior or clinics' capacity - may bias the population captured by surveillance efforts under both strategies. Given the additional effort involved in collecting and submitting cultured isolates, we expect the bias to be stronger for culture-based surveillance. NAATs are the clinical standard of care for gonorrhea diagnosis, and they are performed in much higher volume than gonorrhea culture. We also did not explicitly account for lab processing times under different surveillance methods, as these are highly context-dependent.

Our model considered high-level tetracycline resistance conferred by *tetM*. While it is expected that *tetM* also confers doxycycline resistance, the relationship between tetracycline and doxycycline resistance in the context of doxy-PEP is not yet fully understood. Prior studies of minocycline pre-exposure prophylaxis have shown failure at a tetracycline MIC > 2 ug/mL (5). Culture-based surveillance can identify low-level tetracycline resistance in strains without *tetM* (minimum inhibitory concentrations 2-8 μg/mL) in addition to high-level tetracycline resistance driven by *tetM*. In the US, *tetM* is present in about 10% of all isolates and about a third of tetracycline-resistant isolates (4). However, it is noteworthy that in some locations, *tetM* has been detected in up to 100% of all *N. gonorrhoeae* isolates (6). While assessing for the presence of *tetM* in remnant NAATs will not capture all instances of tetracycline resistance, *tetM* remains a critically important resistance indicator that may become more prevalent with widespread use of doxy-PEP.

**Figure S1. Compartmental model structure**

Deterministic compartmental model with disease states susceptible ($S$), exposed ($E$), symptomatic infectious ($Y$), and asymptomatic infectious ($Z$), with subscripts indicating strains that are antibiotic susceptible ($0$), ceftriaxone resistant ($c$), doxycycline resistant ($d$), and dually ceftriaxone and doxycycline resistant ($cd$).

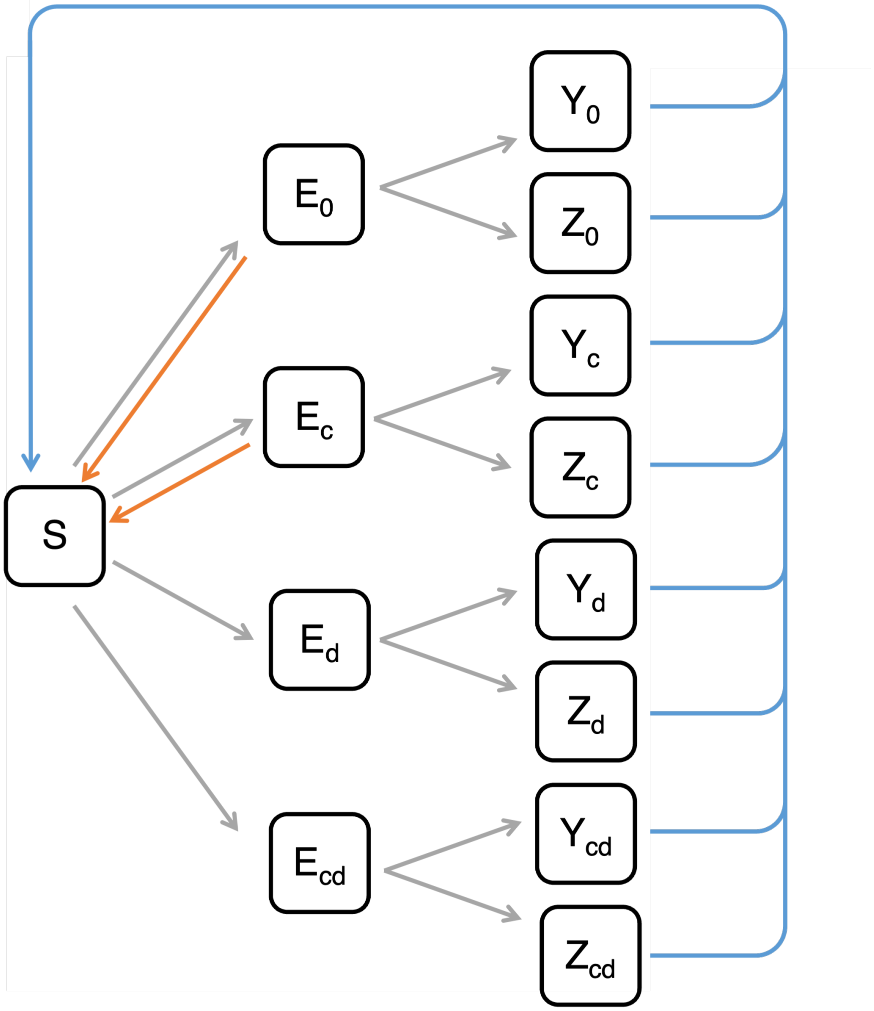

**Model Equations**

*Susceptible compartment*

*dS/dt = population entry – population exit – exposure + natural recovery + treatment (symptomatic + asymptomatic) + retreatment (symptomatic) + doxy-PEP success*

$$\frac{dS}{dt} = \mu_{entry}N - \mu_{exit}S - \frac{bK\left(Y_0 + Z_0 + \phi_c(Y_c + Z_c) + \phi_d(Y_d + Z_d) + \phi_{cd}(Y_{cd} + Z_{cd})\right)}{N}S + \gamma_n(Y_0 + Z_0 + Y_c + Z_c + Y_d + Z_d + Y_{cd} + Z_{cd}) + \theta_s(Y_0 + Y_d) + \theta_a(Z_0 + Z_d) + \theta_e(Y_c + Y_{cd}) + \alpha(1-\rho)(E_0 + E_c))$$

*Exposed compartments*

*dE/dt = - population exit + exposure – infection (symptomatic + asymptomatic)*

$$\frac{dE_0}{dt} = \frac{bK(Y_0 + Z_0)}{N}S - E_0$$

$$\frac{dE_c}{dt} = \frac{bK\phi_c(Y_c + Z_c)}{N}S - E_c$$

$$\frac{dE_d}{dt} = \frac{bK\phi_d(Y_d + Z_d)}{N}S - E_d$$

$$\frac{dE_{cd}}{dt} = \frac{bK\phi_{cd}(Y_{cd} + Z_{cd})}{N}S - E_{cd}$$

*Symptomatic infectious compartments*

*dI/dt = - population exit + infection – natural recovery – treatment – retreatment*

$$\frac{dY_0}{dt} = -\mu_{exit}Y_0 + \sigma(1 - \alpha + \alpha\rho)E_0 - \gamma_n Y_0 - \theta_s Y_0$$

$$\frac{dY_c}{dt} = -\mu_{exit}Y_c + \sigma(1 - \alpha + \alpha\rho)E_c - \gamma_n Y_c - \theta_e Y_c$$

$$\frac{dY_d}{dt} = -\mu_{exit}Y_d + \sigma E_d - \gamma_n Y_d - \theta_s Y_d$$

$$\frac{dY_{cd}}{dt} = -\mu_{exit}Y_{cd} + \sigma E_{cd} - \gamma_n Y_{cd} - \theta_e Y_{cd}$$

*Asymptomatic infectious compartments*

*dI/dt = - population exit + infection – natural recovery – treatment*

$$\frac{dZ_0}{dt} = -\mu_{exit}Z_0 + (1-\sigma)(1-\alpha+\alpha\rho)E_0 - \gamma_n Z_0 - \theta_a Z_0$$

$$\frac{dZ_c}{dt} = -\mu_{exit}Z_c + (1-\sigma)(1-\alpha+\alpha\rho)E_c - \gamma_n Z_c$$

$$\frac{dZ_d}{dt} = -\mu_{exit}Z_d + (1-\sigma)E_d - \gamma_n Z_d - \theta_a Z_d$$

$$\frac{dZ_{cd}}{dt} = -\mu_{exit}Z_{cd} + (1-\sigma)E_{cd} - \gamma_n Z_{cd}$$

**Table S1. Model parameters**
Parameter descriptions and values, adapted from Reichert and Grad (2023) (3).

| Parameter | Description | Value |
|---|---|---|
| $N$ | Population size | $1{,}000{,}000$ |
| $n_1, n_2, n_3$ | Relative size of sexual activity group | Low risk: $0.3$<br>Intermediate risk: $0.6$<br>High risk: $0.1$ |
| | Partner change rate, by sexual activity group, per year | Low risk: $1.41$<br>Intermediate risk: $5 * 1.41$<br>High risk: $20 * 1.41$ |
| $\epsilon$ | Assortative mixing parameter | $0.24$ |
| | Initial ceftriaxone resistance proportion (starting condition) | $0.0001$ |
| | Initial high-level doxycycline resistance proportion (starting condition) | $0.109$ |
| | Initial gonorrhea prevalence (starting condition) | $0.03$ |
| $\mu_{entry}, \mu_{exit}$ | Rates of population entry and exit, per person per year | $1/20$ |
| $b$ | Transmission probability (per partnership) | $0.55$ |
| $K$ | Contact matrix | |
| $\sigma$ | Proportion of incident infections that are symptomatic | $0.45$ |
| $\gamma_n$ | Natural recovery rate, per day | $1 / 76$ |
| $\theta_s$ | Symptomatic care seeking rate, per day | $1 / 15$ |
| $\theta_a$ | Asymptomatic screening rate, per year | $0.36$ |
| $\theta_e$ | Symptomatic retreatment rate (given failure of first treatment), per day | $0.9 * \theta_s/3$ |
| $\phi_c, \phi_d, \phi_{cd}$ | Relative fitness, by strain | ceftriaxone-resistant: $\phi_c = 0.98$<br>doxycycline-resistant: $\phi_d = 0.98$<br>dual resistant: $\phi_{cd} = \phi_c * \phi_d$ |
| $\alpha$ | Doxy-PEP uptake rate (proportion of exposed population) | Varying, $0.1 - 0.9$ |
| $\rho$ | Relative risk of infection when using doxy-PEP | doxycycline-resistant strains: $1$<br>doxycycline-susceptible strains: $0.38$ |
| | Average time exposed | 1 day |

**Figure S2. Delay in 95% confident estimate of rising resistance for varying doxy-PEP uptake levels**

Distributions (25th percentile, median, and 75th percentile) of the time delay until attaining 95% confidence in crossing a resistance threshold ranging from 0.11 to 0.3 relative to the true time it takes for the resistance to reach the threshold, for doxy-PEP uptake rates ranging from 10-90% (shades of pink), comparing culture-based and NAATs-based surveillance with sampling intensities of 25 monthly cultured specimens and 20% of positive NAATs, respectively.

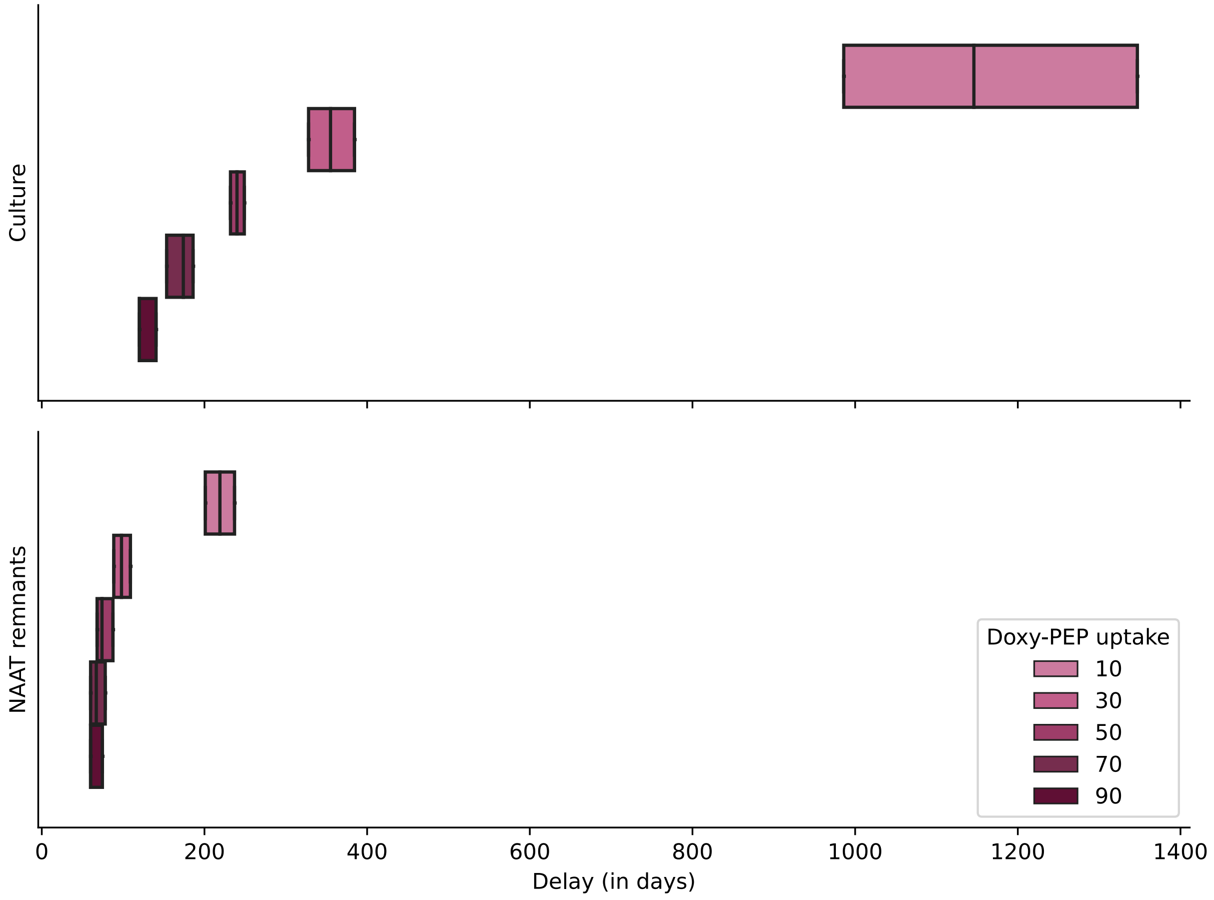
